# Transforming Healthcare Education: Harnessing Large Language Models for Frontline Health Worker Capacity Building using Retrieval-Augmented Generation

**DOI:** 10.1101/2023.12.15.23300009

**Authors:** Yasmina Al Ghadban, Huiqi (Yvonne) Lu, Uday Adavi, Ankita Sharma, Sridevi Gara, Neelanjana Das, Bhaskar Kumar, Renu John, Praveen Devarsetty, Jane E. Hirst

## Abstract

In recent years, large language models (LLMs) have emerged as a transformative force in several domains, including medical education and healthcare. This paper presents a case study on the practical application of using retrieval-augmented generation (RAG) based models for enhancing healthcare education in low- and middle-income countries. The model described in this paper, SMART*health* GPT, stems from the necessity for accessible and locally relevant medical information to aid community health workers in delivering high-quality maternal care. We describe the development process of the complete RAG pipeline, including the creation of a knowledge base of Indian pregnancy-related guidelines, knowledge embedding retrieval, parameter selection and optimization, and answer generation. This case study highlights the potential of LLMs in building frontline healthcare worker capacity and enhancing guideline-based health education; and offers insights for similar applications in resource-limited settings. It serves as a reference for machine learning scientists, educators, healthcare professionals, and policymakers aiming to harness the power of LLMs for substantial educational improvement.

## 1 Introduction

Recently, the natural language processing (NLP) landscape has seen spectacular advances with the increasing availability of pre-trained large language models (LLMs), such as Open AI’s GPT [1], Llama [2], and PaLM [3]. These models have had applications in several fields and are increasingly being employed in medical education and healthcare [4, 5].

Retrieval-Augmented Generation (RAG) and fine-tuning emerge as two powerful methodologies for tailoring pre-trained LLMs to specific applications. Fine-tuning modifies the model’s weight based on a task-specific dataset in a “close-book” setting, relying solely on additional input-output pairs of training data for learning [6, 7]. In contrast, RAG operates in an “open-book” setting and does not require labelled training data [8, 9]. Instead, it utilizes external information sources to retrieve and incorporate relevant information, enhancing the model’s comprehension and generative capabilities.

This paper introduces a case study, SMART*health* GPT (version Rv1), that showcases the application of RAG in developing educational and communication tools for frontline healthcare workers in low- and middle-income countries. This LLM tool aims to enhance community health workers’ knowledge, skills, and competencies by providing accessible, context-relevant information.

## 2 SMART*health* Pregnancy

Identifying women with high-risk pregnancies before complications occur is essential to prevent maternal and newborn mortality and morbidity. However, owing to health worker shortages, resource constraints, poverty, and gender barriers, delivering high-quality pregnancy and postnatal care to women living in rural locations in low- and middle-income countries is challenging.

To improve the early detection, referral and management of high-risk pregnancy conditions and early prevention of non-communicable diseases, SMART*health* Pregnancy, a digitally supported tool, was developed for frontline health workers (ASHAs) [10, 11]. The system utilises task sharing to ASHAs, equipped with point-of-care devices and an Android tablet App with electronic decision support based on the George Institute for Global Health (TGI) SMART*health* platform. The system has been co-designed with end users in rural India, demonstrated to be feasible and acceptable, and is currently being assessed in a large cluster implementation effectiveness trial across 60 villages in two states in India (Telangana and Haryana) (clinicaltrial.gov NCT05752955).

Qualitative research has found that whilst the system is highly valued, ASHAs lack detailed knowledge to give diet and lifestyle advice, and information about medical conditions and pregnancy symptoms in simple terms and in local languages to support pregnant and postpartum women. In line with this need, the aim of this project was to develop, technically and clinically validate, an LLM suitable for community health workers in rural India to support guideline-based pregnancy care. We believe this LLM application, SMART*health* GPT, can be a valuable tool to enhance medical education for frontline health workers, particularly in resource-constrained environments.

## 3 Methodology

### 3.1 Formal definition of RAG

RAG enables LLMs to access information from non-parametric storage, making it highly adaptable to new tasks and reducing the need for extensive annotated training data. Within the RAG framework, external information sources are transformed into embeddings and stored in a vector database. This process forms the basis for the subsequent steps. To illustrate the RAG process formally, we express it using conditional probabilities, dividing it into two key components: retrieval and generation.

In the retrieval step, the objective is to select a set of relevant documents, *D*(*D*_1_,…, *D*_*k*_), from a repository of documents (*E*) based on a user’s question (*Q*). This can be expressed as *P* (*D*|*Q*), the likelihood of choosing documents D from E, given the question *Q*. This probability is computed by the retriever.

Once the relevant documents (*D*) have been retrieved, the answer (*A*) needs to be generated. This generation step can be expressed as *P* (*A*|*D, Q*), representing the probability distribution of generating the answer *A*, given the retrieved documents *D* and the question *Q*. This probability is computed by a text generator.

The entire RAG process can be expressed as follows:

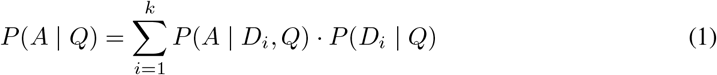

Here, *P* (*A*|*Q*) signifies the probability of generating answer A given the question Q.

In summary, RAG selects pertinent documents based on the input context and then generates the output text, conditioned on both the selected documents and the input context. These steps are often implemented using neural network models, which are trained to maximize the likelihood of generating the correct output, aligning with the retrieved documents.

### 3.2 Rationale for the use of RAG

We selected RAG for our use case for several reasons. First, the pedagogical nature of our application necessitates that responses provided by the model are not only accurate but also traceable back to their sources. RAG’s ability to trace responses back to their respective sources increases explainability and trustworthiness in the educational content and reduces potential model hallucinations. This also aligns with the needs of our application where ASHAs not only require a response to their question but also a source guideline to refer to. The RAG method allows us to retrieve the context relevant to the query and to return the source (document name and page number) to the user. Second, our model needs to be scalable, especially in the context of vast knowledge bases within the healthcare domain. RAG models are highly scalable as they leverage retrieval mechanisms to accommodate large knowledge bases. Third, given the continuous evolvement of clinical guidelines, our model needs to be flexible and swiftly updated to align with the latest recommendations. By using RAG, our model can be easily and quickly updated by incorporating new guidelines or updates into the knowledge base, ensuring the relevance and accuracy of our model.

### 3.3 Model development

We deployed a three-step approach in the RAG model development pipeline, as shown in Figure 1.

**Figure 1.**
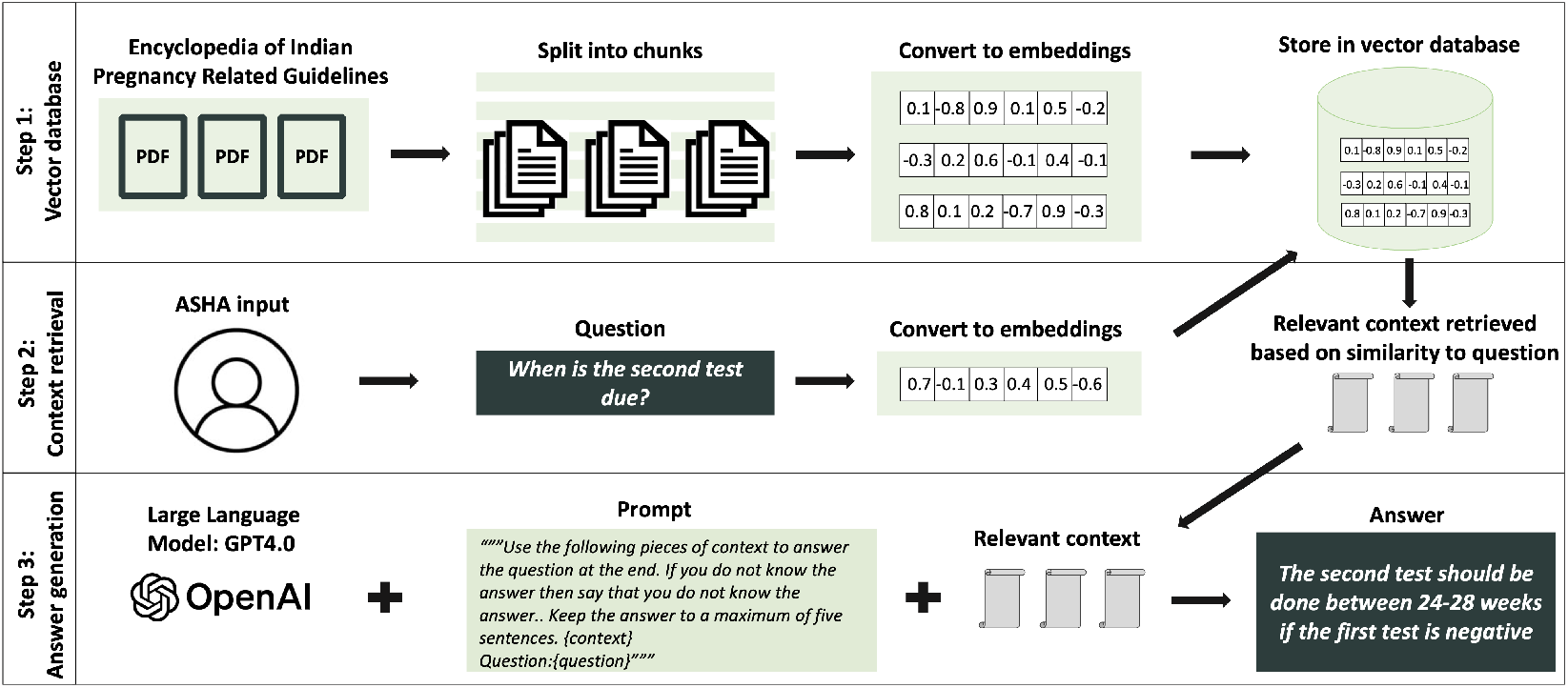
Flow diagram of the RAG process in SMART*health* GPT

**Figure 2.**
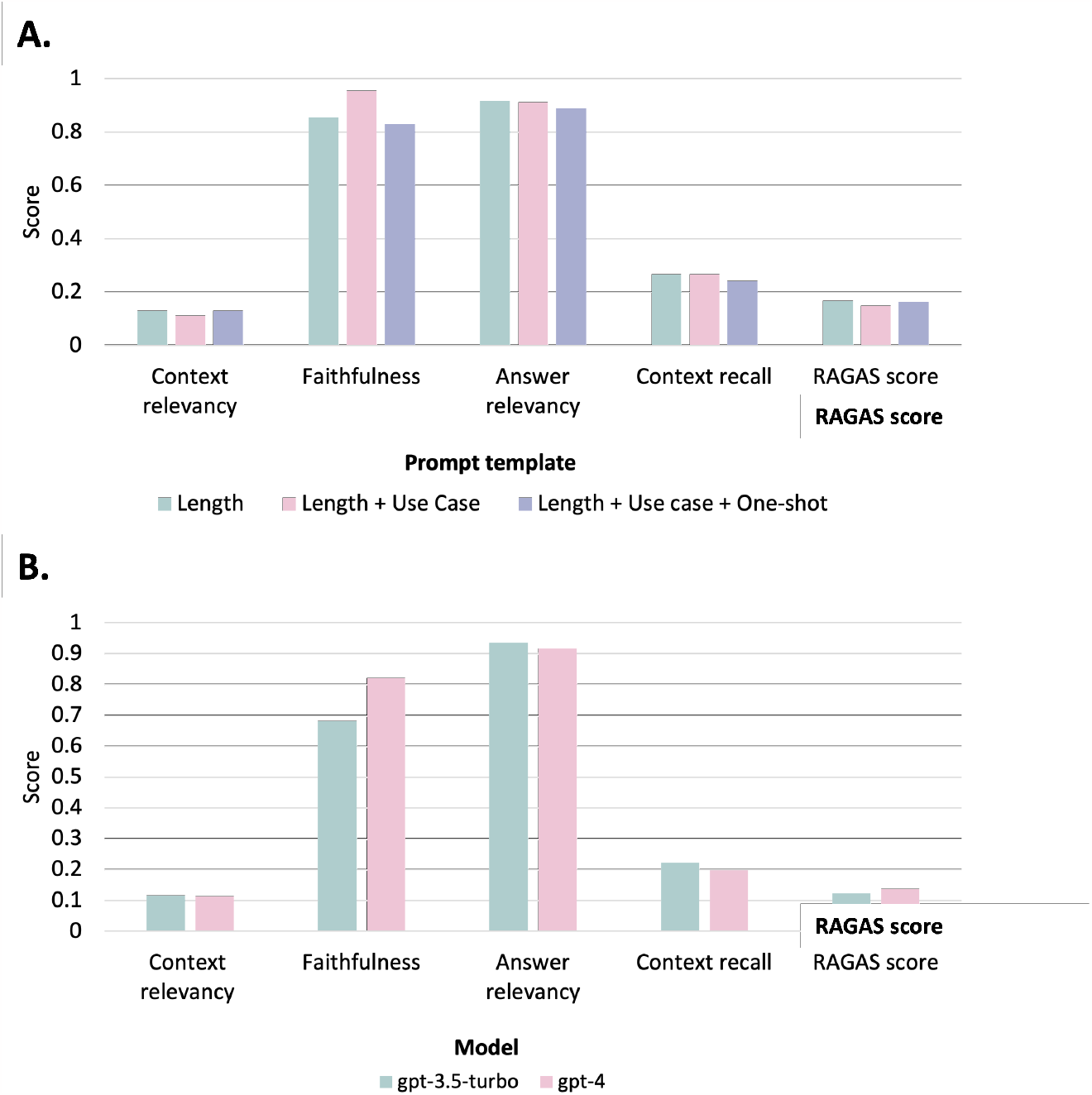
RAGAS metrics for different model parameters. (A) Prompt template (B) Model type

#### Step 1: Development of the encyclopaedia of Indian guidelines

To create the knowledge base that RAG will be based on, the clinical team created a large repository of Indian pregnancy-related guidelines. This initial file included 37 documents. The clinical team then created a smaller repository of Indian pregnancy guidelines relevant only to community health workers, ASHAs. To identify whether the repository was complete, we first ran the model of 130 questions collected through community engagement with ASHAs on the small repository. For questions where the model did not return an answer, we ran the model on the large repository and identified the sources that were missing. The clinical team then decided whether the source should be included or if it was appropriate for the model not to return an answer to that question.

We transformed the final repository of Indian guidelines into embeddings and stored them in the same FAISS vector database. We chose the FAISS vector store for its ability to perform similarity search in sets of vectors of any size, and the option to store the index locally. This allows us to load the vector store directly, rather than creating it at every iteration, decreasing the processing time.

#### Step 2.a: Context retrieval

We selected three commonly used retrieval methods in RAG: vector-store backed retriever, contextual compression retriever and ensemble retriever, as shown in Supplementary Figure S1. The vector-store backed retriever is the simplest method, which retrieves documents with the highest similarity to the question.

In this study, all three retriever methods were tested and compared for both similarity search and Maximal Marginal Relevance (MMR) search. Compared to the similarity search, MMR balances between relevance and diversity. It selects the document *D*_*i*_ from the ranked list of relevant documents, *D*(*D*_1_,…, *D*_*k*_), that maximizes the trade-off between similarity to question Q, *Sim*_1_(*D*_*i*_, *Q*) and similarity to the documents already selected (S), *Sim*_2_(*D*_*i*_, *D*_*j*_) [12]. A parameter, *\* controls the trade off between *Sim*_1_(*D*_*i*_, *Q*) and *Sim*_2_(*D*_*i*_, *D*_*j*_). MMR can be defined as:

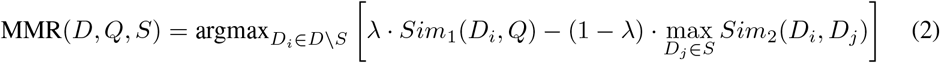

Similarity is measured using the cosine similarity between two vectors **A** and **B**, which is calculated as follows:

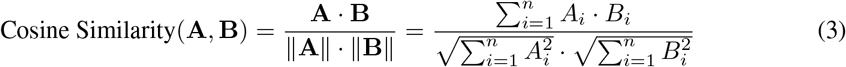

The selection of the retrieval method was based on a quantitative assessment and an evaluation of the processing time (using both wall time and CPU time). For the quantitative assessment, we used 10 diverse pregnancy-related questions and compared the model’s answer to the clinician provided answer (the gold standard) using both a cosine similarity score and a Clinical BERT similarity score. ClinicalBERT is a modified BERT model pre-trained on patient clinical notes and electronic health records, which more accurately captures clinical word similarity [13]. Given the clinical nature of our application, we used a similarity score based on the ClinicalBERT model (range 0-5) in addition to the cosine similarity score (range 0-1).

#### Step 2.b: Parameter optimisation

We then investigated the impact of 4 model parameters (chunk size, chunk overlap, number of documents retrieved (k) and search type) on model performance. In this round, we limited our retrieval method to a vector-store backed retrieval but compared similarity search and MMR. Due to the limited number of questions, we carried out two grid search tests on 1) chunk size and chunk overlap, and 2) search type and number of documents retrieved (k).

To select the model parameters, we developed an evaluation pipeline using the RAGAS framework, which allows evaluation of both generation and retrieval steps alone, and a RAGAS score for overall performance assessment [14]. The metrics that evaluate retrieval are **context relevancy** which measures the signal-to-noise ratio in retrieved contexts by determining the ratio of essential sentences to total sentences in the context, and **context recall** which assesses the retriever’s ability to find all necessary information by checking if statements from the ground-truth answer are present in the retrieved context. The metrics that evaluate generation are **faithfulness** which assesses the factual accuracy of the generated answer by comparing its statements to the context and calculating the ratio of correct statements to total statements, and **answer relevancy** which measures how relevant the generated answer is to the original question by assessing its similarity to potential questions it could address. The RAGAS score is the harmonic mean of all four metrics [14].

#### Step 3: Answer generation

The backbone of the SMART*health* GPT Rv1 is the Open AI GPT model. To optimise the answer generation, we compared two pre-trained models: “gpt-3.5-turbo” and “gpt-4”. In this answer generation step, we also designed three prompt instructions that were customised to meet clinical needs, namely the length of response; length and use case; and length and use case and one-shot learning. We evaluated model performance based on the RAGAS metrics.

## 4 Model performance evaluation and results

### Step 1: Development of the encyclopaedia of Indian guidelines

The final repository included 20 pregnancy-related guidelines. The included guidelines covered a range of pregnancy-related conditions with a particular focus on the three key conditions addressed by the SMART*health* Pregnancy project: anaemia, gestational diabetes, and hypertension in pregnancy.

### Step 2.a: Context retrieval

The answer quality (measured by ClinicalBERT and cosine similarity scores) of the simplest vector-store based retriever did not improve with the contextual compression retriever or the ensemble retriever, while the processing time increased, as shown in Supplementary Table S1. The significant response time increase was deemed not feasible for our application, considering the time pressure ASHAs face while providing care for women. Therefore, the simplest vector-store based retriever was chosen to be evaluated in the next steps.

### Step 2.b: Parameter optimisation

Interestingly, the chunk size, chunk overlap, search type and number of retrieved documents (k) parameters did not significantly impact model performance, measured by RAGAS metrics (Table 1 and Supplementary Table S2). As a result, model parameters were primarily chosen based on processing time and technical considerations. Accordingly, we selected a chunk size of 1000, with an overlap of 200 characters. We chose the MMR search type as health guidelines include several similar parts, and considering diversity in retrieval minimises repetition and provides more comprehensive documents. Two retrieved documents (k=2) showed a balance in result completeness and duplications.

**Table 1:** RAGAS metrics for SMART*health* GPT model parameters: search type and number of retrieved documents (k)

| k | Evaluation metric |  |  |  |  |
| --- | --- | --- | --- | --- | --- |
|  | Context relevancy | Faithfulness | Answer relevancy | Context recall | RAGAS score |
| MMR search |  |  |  |  |  |
| 1 | 0.16 | 0.77 | 0.91 | 0.26 | 0.15 |
| 2 | 0.12 | 0.85 | 0.92 | 0.22 | 0.15 |
| 3 | 0.08 | 0.93 | 0.92 | 0.14 | 0.11 |
| 4 | 0.07 | 0.89 | 0.91 | 0.31 | 0.13 |
| 5 | 0.07 | 0.85 | 0.93 | 0.24 | 0.10 |
| Similarity search |  |  |  |  |  |
| 1 | 0.10 | 0.72 | 0.93 | 0.28 | 0.17 |
| 2 | 0.09 | 0.86 | 0.91 | 0.21 | 0.12 |
| 3 | 0.10 | 0.90 | 0.91 | 0.37 | 0.20 |
| 4 | 0.10 | 0.81 | 0.91 | 0.19 | 0.12 |
| 5 | 0.10 | 0.90 | 0.91 | 0.40 | 0.20 |

### Step 3: Answer generation

The model choice and prompt template also minimally impacted RAGAS metrics. We selected “gpt-4” due to its lower likelihood of hallucinations as demonstrated in previous studies [15], and the one-shot prompt template as providing an example may result in more predictable responses, required for our application.

## 5 Initial clinical validation results

The model is currently undergoing a first round of clinical validation. During this round, 12 community medicine clinicians and two obstetricians, across two states, rated the model generated answers based on accuracy, completeness, appropriateness, and presence of bias on a 3-point Likert scale. The 180 questions included in this round of clinical validation were developed with ASHAs directly though focus groups and community engagement. Each question was rated by a different number of clinicians between two and six. For 141 (79%) questions, all clinicians agreed that the AI generated answer was completely or partially accurate; and that the AI-generated answer was either unbiased or actively promoted equity. However, all clinicians rated the completeness of the AI generated answer as adequate or comprehensive for only 49 (35%) questions. The clinical validation has allowed to gain significant insight into the performance of the model, and importantly, has allowed us to identify the cases where the model is failing (the answers with low ratings).

## 6 Future work

Our immediate next goal is the improvement of responses using prompt engineering and ensuring the LLM includes safety features, such as moderation, bias and evaluation checks. In collaboration with the clinical team, we also need to define topics that are beyond the scope of community health workers, such as questions around medical prescriptions and interpretation of reports. Additionally, we need to develop contextually relevant and gender transformative responses to sensitive queries such as questions about predicting the gender of the baby.

In addition to rating the model’s answer to the 180 questions, clinicians also provided an ideal response that is clear, uncontroversial, and appropriate for ASHAs. The answers provided by multiple clinicians are being compiled and standardized by an obstetrician and a community medicine doctor.

These answers will constitute the “gold-standard” responses and the repository of 180 question-answer pairs can be used to fine-tune the model or improve its prompts. The LLM will be improved iteratively based on the feedback from each phase of validation (Figure 3).

**Figure 3.**
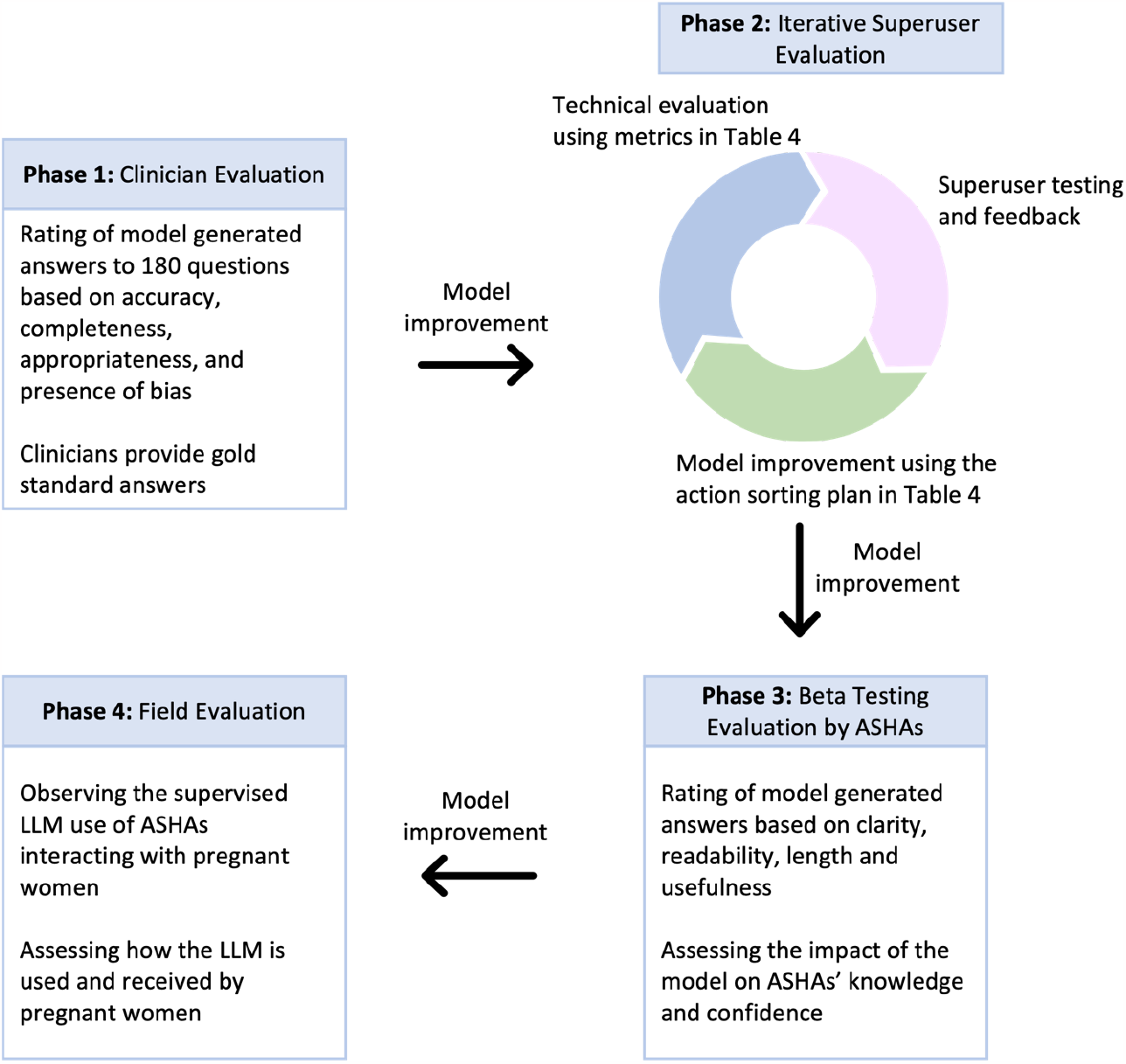
Iterative LLM improvement through the four phases of clinical evaluation

Future work must also address several impending challenges. These include scaling up the knowledge base while ensuring efficient information retrieval, handling multi-level conversations, as well as addressing critical data privacy concerns, such as communication storage management and learning from user requests. A future avenue of work is the exploration of local models to overcome the limitations imposed by the use of Open-AI models such as cost.

## 7 Conclusion

In this paper, we have described a comprehensive case study centred on the deployment of RAG to develop SMART*health* GPT (version Rv1), an LLM tool to build the capacity of community health workers. This model was developed to enhance guideline-based pregnancy care, showcasing the impactful intersection of generative AI and healthcare education. The selection of RAG was driven by clinical needs – traceability to source material, scalability across vast knowledge bases, and seamless adaptability to evolving clinical guidelines.

Our process of model development and optimization encompassed the building of the encyclopaedia of Indian pregnancy-related guidelines and quantitative evaluations of different retriever methods and model parameters. SMART*health* GPT showcases the promising role of RAG and LLMs in medical education and provides insights for future applications of generative AI in diverse educational settings. We hope that the deployment of RAG models emerges as a strategy for surmounting educational barriers, and the case study of SMART*health* GPT (version Rv1) can serve as a template for the application of generative AI in education, particularly in non-traditional educational settings and resource-constrained environments.

## Data Availability

All data produced in the present study are available upon reasonable request to the authors

## Acknowledgments and Disclosure of Funding

This study is funded by the Bill and Melinda Gates Foundation Grand Challenges Equitable AI. The views expressed are those of the authors and not necessarily those of BMGF. The funder has no role in the study design, data collection, management, analysis and interpretation; writing or the report; and decisions related to publication and presentation of findings. The authors declare no financial interests.

## References

[1] Tom B. Brown et al. Language Models are Few-Shot Learners. arXiv:2005.14165 [cs]. July 2020. DOI: 10.48550/arXiv.2005.14165. URL: http://arxiv.org/abs/2005.14165 (visited on 09/25/2023).

[2] Hugo Touvron et al. LLaMA: Open and Efficient Foundation Language Models. arXiv:2302.13971 [cs]. Feb. 2023. DOI: 10.48550/arXiv.2302.13971. URL: http://arxiv.org/abs/2302.13971 (visited on 09/25/2023).

[3] Aakanksha Chowdhery et al. PaLM: Scaling Language Modeling with Pathways. arXiv:2204.02311 [cs]. Oct. 2022. DOI: 10.48550/arXiv.2204.02311. URL: http://arxiv.org/abs/2204.02311 (visited on 09/25/2023).

[4] Rachel S. Goodman et al. “On the cusp: Considering the impact of artificial intelligence language models in healthcare”. English. In: Med 4.3 (Mar. 2023). Publisher: Elsevier, pp. 139–140. ISSN: 2666-6359, 2666-6340. DOI: 10.1016/j.medj.2023.02.008. URL: https://www.cell.com/med/abstract/S2666-6340(23)00068-5 (visited on 08/30/2023).

[5] Conrad W. Safranek et al. “The Role of Large Language Models in Medical Education: Applications and Implications”. EN. In: JMIR Medical Education 9.1 (Aug. 2023). Company: JMIR Medical Education Distributor: JMIR Medical Education Institution: JMIR Medical Education Label: JMIR Medical Education Publisher: JMIR Publications Inc., Toronto, Canada, e50945. DOI: 10.2196/50945. URL: https://mededu.jmir.org/2023/1/e50945 (visited on 09/23/2023).

[6] Hyung Won Chung et al. Scaling Instruction-Finetuned Language Models. arXiv:2210.11416 [cs]. Dec. 2022. DOI: 10.48550/arXiv.2210.11416. URL: http://arxiv.org/abs/2210.11416 (visited on 09/25/2023).

[7] Jason Wei et al. Finetuned Language Models Are Zero-Shot Learners. arXiv:2109.01652 [cs]. Feb. 2022. DOI: 10.48550/arXiv.2109.01652. URL: http://arxiv.org/abs/2109.01652 (visited on 09/25/2023).

[8] Matthew Lamm et al. “QED: A Framework and Dataset for Explanations in Question Answering”. In: Transactions of the Association for Computational Linguistics 9 (2021). Place: Cambridge, MA Publisher: MIT Press, pp. 790–806. DOI: 10.1162/tacl_a_00398. URL: https://aclanthology.org/2021.tacl-1.48 (visited on 09/25/2023).

[9] Patrick Lewis et al. Retrieval-Augmented Generation for Knowledge-Intensive NLP Tasks. arXiv:2005.11401 [cs]. Apr. 2021. DOI: 10.48550/arXiv.2005.11401. URL: http://arxiv.org/abs/2005.11401 (visited on 09/25/2023).

[10] Shobhana Nagraj et al. “SMARThealth Pregnancy: Feasibility and Acceptability of a Complex Intervention for High-Risk Pregnant Women in Rural India: Protocol for a Pilot Cluster Randomised Controlled Trial”. In: Frontiers in Global Women’s Health 2 (2021). ISSN: 2673-5059. URL: https://www.frontiersin.org/articles/10.3389/fgwh.2021.620759 (visited on 09/21/2023).

[11] Shobhana Nagraj et al. “A Mobile Clinical Decision Support System for High-Risk Pregnant Women in Rural India (SMARThealth Pregnancy): Pilot Cluster Randomized Controlled Trial”. In: JMIR Formative Research 7 (July 2023), e44362. ISSN: 2561-326X. DOI: 10.2196/44362. URL: https://www.ncbi.nlm.nih.gov/pmc/articles/PMC10401191/ (visited on 09/25/2023).

[12] Jade Goldstein and Jaime Carbonell. “Summarization: (1) Using MMR for Diversity-Based Reranking and (2) Evaluating Summaries”. In: TIPSTER TEXT PROGRAM PHASE III: Proceedings of a Workshop held at Baltimore, Maryland, October 13–15, 1998. Baltimore, Maryland, USA: Association for Computational Linguistics, Oct. 1998, pp. 181–195. DOI: 10.3115/1119089.1119120. URL: https://aclanthology.org/X98-1025 (visited on 09/25/2023).

[13] Kexin Huang, Jaan Altosaar, and Rajesh Ranganath. ClinicalBERT: Modeling Clinical Notes and Predicting Hospital Readmission. arXiv:1904.05342 [cs]. Nov. 2020. URL: http://arxiv.org/abs/1904.05342 (visited on 09/25/2023).

[14] Evaluating RAG pipelines with Ragas + LangSmith. en. Aug. 2023. URL: https://blog.langchain.dev/evaluating-rag-pipelines-with-ragas-langsmith/ (visited on 09/21/2023).

[15] Rohaid Ali et al. “Performance of ChatGPT, GPT-4, and Google Bard on a Neurosurgery Oral Boards Preparation Question Bank”. eng. In: Neurosurgery (June 2023). ISSN: 1524-4040. DOI: 10.1227/neu.0000000000002551.

[16] Gordon V. Cormack, Charles L A Clarke, and Stefan Buettcher. “Reciprocal rank fusion outperforms condorcet and individual rank learning methods”. In: Proceedings of the 32nd international ACM SIGIR conference on Research and development in information retrieval. SIGIR ‘09. New York, NY, USA: Association for Computing Machinery, July 2009, pp. 758–759. ISBN: 978-1-60558-483-6. DOI: 10.1145/1571941.1572114. URL: https://dl.acm.org/doi/10.1145/1571941.1572114 (visited on 09/25/2023).

