## Supplemental Material for "Transforming Healthcare Education: Harnessing Large Language Models for Frontline Health Worker Capacity Building using Retrieval-Augmented Generation"

### 8 Supplementary Material

The vector-store backed retriever is the simplest method, which retrieves documents with the highest similarity to the question. The contextual compression retriever first retrieves documents using a base-retriever (such as a vector-store backed retriever) and then compresses the contents of the retrieved documents. This method increases the number of results we can pass to the LLM by shrinking the results to only the most relevant information. The ensemble retrieval combines multiple retrievers and re-ranks the results based on the reciprocal ranking algorithm [16]. This method leverages the strengths of different retrieval methods. We combined a vector-based retriever with a sparse retriever (BM25). The vector-based retriever finds relevant documents based on similarity, while the sparse retriever finds relevant documents based on keywords. The BM25 uses the term frequency of each term in both the query and the documents, as well as the documents' length to compute a score for each document in response to the query.

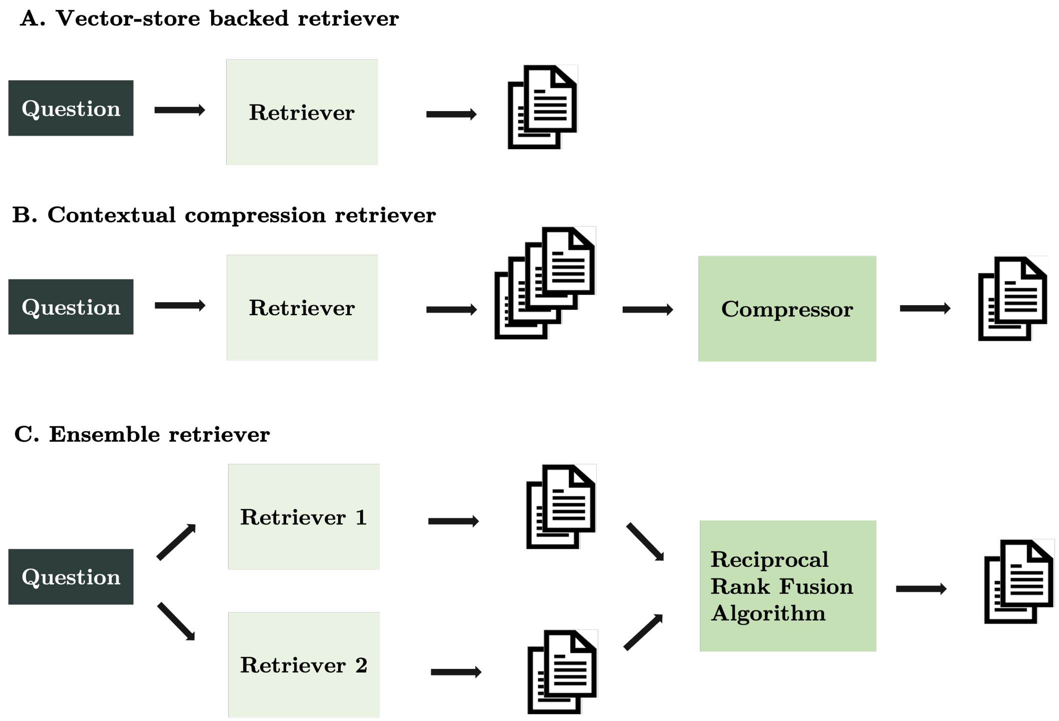


Figure S1: Retrieval mechanisms considered in model development (A) Vector-store backed retriever (B) Contextual compression retriever (C) Ensemble retriever

Table S1: Processing time (s) and similarity score results for different retrieval methods

| Search type | Processing time (s) | Similarity score | ClinicalBERT similarity score |
|---|---|---|---|
| **Vector-store backed retriever** | | | |
| Similarity | 12.5 | 0.89 | 2.4 |
| MMR | 12.8 | 0.89 | 2.4 |
| **Contextual compression retriever** | | | |
| Similarity | 18.9 | 0.9 | 2.6 |
| MMR | 18.9 | 0.89 | 2.5 |
| **Ensemble retriever** | | | |
| Similarity | 14.6 | 0.89 | 2.4 |
| MMR | 15.3 | 0.88 | 2.5 |

Table S2: RAGAS metrics for SMART*health* GPT model parameters: chunk size and chunk overlap (in characters)

| Size (overlap) | Evaluation metric | | | | |
|---|---|---|---|---|---|
| | Context relevancy | Faithfulness | Answer relevancy | Context recall | RAGAS score |
| 500 (100) | 0.13 | 0.87 | 0.92 | 0.13 | 0.12 |
| 500 (200) | 0.08 | 0.79 | 0.92 | 0.11 | 0.07 |
| 1000 (100) | 0.08 | 0.82 | 0.91 | 0.22 | 0.12 |
| 1000 (200) | 0.12 | 0.77 | 0.90 | 0.22 | 0.15 |
| 1000 (500) | 0.13 | 0.80 | 0.91 | 0.12 | 0.12 |
| 2000 (100) | 0.05 | 0.90 | 0.92 | 0.15 | 0.07 |
| 2000 (200) | 0.05 | 0.81 | 0.90 | 0.26 | 0.13 |
| 2000 (500) | 0.07 | 0.98 | 0.90 | 0.21 | 0.11 |
